# Epidemiologic Method Review at Scale: Assessing Charlson Comorbidity Versioning Using a Large Language Model

**DOI:** 10.1101/2025.09.23.25336010

**Authors:** Joshua T. Fuchs, Cara Johnson, Nathan Foster, Peter J. Leese

## Abstract

The Charlson Comorbidity Index (CCI) is widely used in epidemiologic studies. However, many versions of the CCI have been developed since the original method was published in 1987, and it is unclear which version is used most frequently and how version utilization in research has changed over time. We present an approach using a large language model to extract data from articles by detecting which specific CCI version is employed. We analyzed 31,767 articles published since 2012 to evaluate the landscape of CCI implementation. We show that 63% of articles that cite a single method version cite only the original CCI publication, which cannot be applied in the modern real-world data era, leading to ambiguity in actual CCI implementation. More generally, this paper introduces a generalizable approach to scaling methods literature review beyond typical human-review, which we validate and demonstrate through application to the CCI.

## Introduction

The Charlson Comorbidity Index (CCI) was introduced in Charlson et al. 1987^1^ as a quantification of comorbidity, developed and validated on breast cancer patients aiming to predict 1- and 10-year morbidity rates. It provided 19 categories of diseases, assigned each disease category a weight, and generated a single comorbidity index by summing the weights of diseases that were present in a patient.

As demonstrated by the high volume of citations (anywhere from 35k to 65k depending on literature repository) this paper was a landmark study which introduced both a foundational method as well as formalizing a way of thinking about comorbidities that would lead to a proliferation of diverse risk scores for research and clinical care. The CCI has now been widely adopted and validated for use in clinical, epidemiologic, and public health studies as a general indicator of comorbidity burden at an individual and population level^2–5^, and there have been many major and minor CCI methods developments since 1987. We categorize ‘major developments’ as papers that revised or updated the CCI with an intent to change the method for broad, general populations, such as modifying the weights across all the comorbidity categories or updating definitions from ICD-9 to ICD-10 codes^6–11^. These updates differ compared to ‘minor developments’ in the method, such as narrowing the CCI to focus explicitly on a sub-population of patients, or slightly modifying a CCI element such as the diagnosis codes used or adding age as a parameter^12–14^.

While there have been numerous reviews exploring the reliability of versions of the CCI for various clinical outcomes^15^, and studies on methodological application to epidemiologic studies^16–18^, there has not been an analysis across the composite literature that evaluates which method is most frequently implemented, how these method trends change over time, or how often a study is not reproducible because the CCI method employed is unknown. This type of evaluation has been limited by the sheer popularity of including the CCI in studies (over 40,000 articles appear in PubMed Central with the term Charlson Comorbidity Index) as well as the time needed to manually review even a small portion of those articles. In the current study we have used a novel large language model (LLM)-assisted data extraction method to characterize the utilization and evolution of the CCI since 2012 to identify trends and to use as the basis for future CCI methods development.

The automated extraction of data from unstructured text has been shown and utilized in a variety of fields using text mining, natural language processing, and machine learning^19–22^. Recently, there have been demonstrations that LLMs can accurately extract text from scientific texts^23–25^ and can assist in systematic reviews^26^. These text analysis method advancements facilitate significantly larger literature reviews and literature-based analysis than previously when constrained by the feasibility of manually reviewing and extracting information from articles.

Here we present an approach to quantifying the current field of literature for use of the CCI by using a LLM to carefully extract information from available articles. We used a corpus of 31,767 published research articles since 2012 that included the words “Charlson Comorbidity Index”, then systematically analyzed the text using a LLM to identify which references were used for the calculation and implementation of the CCI in that article. The use of the LLM allows us to present a model that goes beyond simple keyword searching or DOI matching, to one that detects nuance in the varied language used to describe methodological implementations. This work allows us to quantify a large number of articles and provides a framework for literature analysis using LLMs.

## Methods

### CCI Version Selection

To quantify the utilization of different CCI methods papers, we selected the ten CCI versions listed in Table 1. These are the most highly-cited methods papers we are aware of, and all have contributed substantially to the development and utilization of the CCI in major and minor ways. We explored the prevalence and utilization of these ten CCI versions in our analysis.

**Table 1:** List of CCI version papers that were included in the analysis. Total citations were collected from SCOPUS.

| CCI Version Author and Year | Total Citations |
| --- | --- |
| Charlson et al. (1987) | 42,035 |
| Deyo, Cherkin, and Ciol (1992) | 9,376 |
| Romano, Roos, and Jollis (1993) | 1,629 |
| Charlson et al. (1994) | 5,624 |
| Klabunde et al. (2000) | 1,690 |
| Halfon et al. (2002) | 200 |
| Schneeweiss et al. (2003) | 275 |
| Sundararajan et al. (2004) | 1,951 |
| Quan et al. (2005) | 8,804 |
| Quan et al. (2011) | 4,518 |

### Article Selection

Figure 1 provides an overview of the selection and pre-processing of articles for analysis by the LLM. Articles were selected from the PubMed Central Open Access Subset^32^, a subset of articles available on PubMed Central for text mining. We selected articles that included the words “Charlson Comorbidity Index” in the title, abstract, or body text. Eligible articles were published between 2012/01/01 and 2025/02/24. The start date for inclusion was the year after the latest CCI version paper was published, see Table 1. This limited a bias where earlier methods had more years to be included in citations and allowed our study to focus on what contemporary researchers have been implementing. We excluded articles that had the word “Review” in the title to focus on articles that calculated the CCI for use in that article. This search yielded 31,767 articles for further analysis.

**Figure 1:**
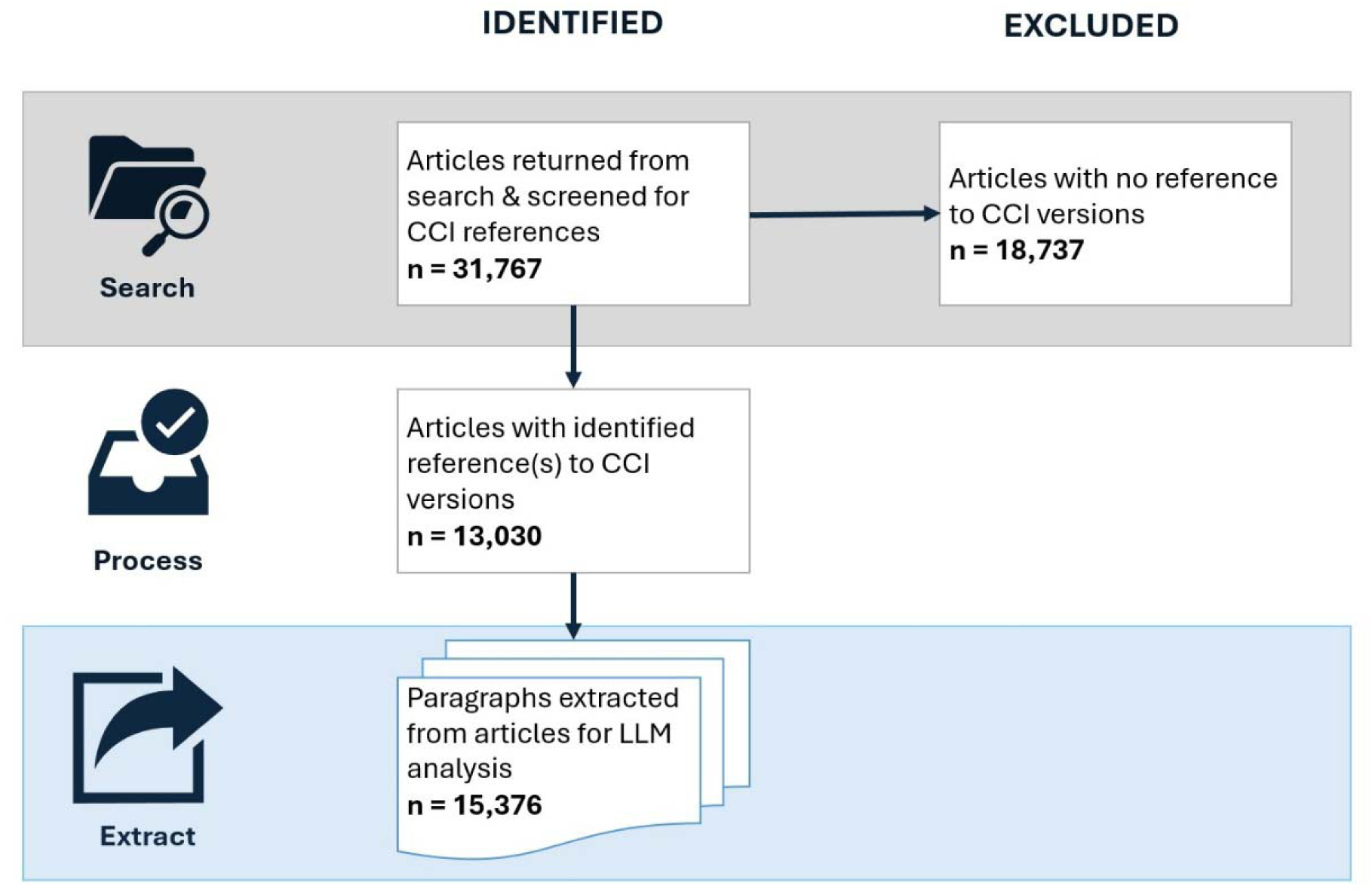
Flow chart of articles from selection in PubMed Central, through preprocessing, and to analysis with the LLM. From 31,767 articles we extracted 15,376 relevant paragraphs that were put into the LLM for data extraction.

Using Bibliometrix^33^, we determined the articles were retrieved from 2,376 unique sources with 126,259 unique authors. The three most common publication types were Journal Articles, Observational Studies, and Clinical Trials, which account for over 88% of the articles. This indicates we collected articles for analysis covering a wide range of publication types and authors, and that our results meaningfully reflect the utilization of the CCI in the research community.

### Article Processing

We extracted the full-text of articles using the Entrez Programming Utilities from the National Center for Biotechnology Information, as implemented in the Biopython package^34,35^. Articles were extracted from PubMed Central as eXtensible Markup Language (XML) files. We parsed the XML file to identify the CCI versions listed in Table 1, then replaced numerical references in the text with the form of [Author Year]. Next, using the pubmed_parser package^36^, we split the text into paragraphs, keeping only the paragraphs that contained references to the CCI versions listed in Table 1. In total, 58.9% (18,737) of articles did not reference one of the CCI versions we searched for and so did not proceed to the data extraction phase. Of the 13,030 articles that did cite one of the CCI versions in Table 1, 87.7% articles had a single paragraph retained for analysis and 12.3% had multiple paragraphs retained. We analyzed only these paragraphs because (1) the details of how the CCI was calculated should nearly always be in paragraphs that include references to the version used, and (2) analyzing the whole article text would contain a significant amount of irrelevant text for the LLM to parse. Analyzing single paragraphs at a time also mitigated having to segment the analysis based on the max token size the model allowed. We analyzed a total of 15,376 paragraphs.

### LLM Data Extraction

Data was extracted from the paragraphs using the Llama 3.3 70B Instruct model^37^. We followed the method of data extraction introduced in Polak & Morgan (2024)^24^ and more generally discussed in Li et al. (2023)^38^. This involved a series of carefully engineered question prompts to extract references that were used to calculate the CCI in that article, if any. To generate the prompts, we went through a standard process of prompt engineering, which has been shown to increase the accuracy of LLM responses^39^.

Figure 2 shows the abbreviated prompts and LLM data extraction flow; full prompts can be found in Appendix A. Two initial questions evaluated if the CCI was calculated in the article (as opposed to solely being referenced) then if the text contained a reference or description of how the CCI was calculated. Negative responses to either of these led to the paragraph not being analyzed further.

**Figure 2:**
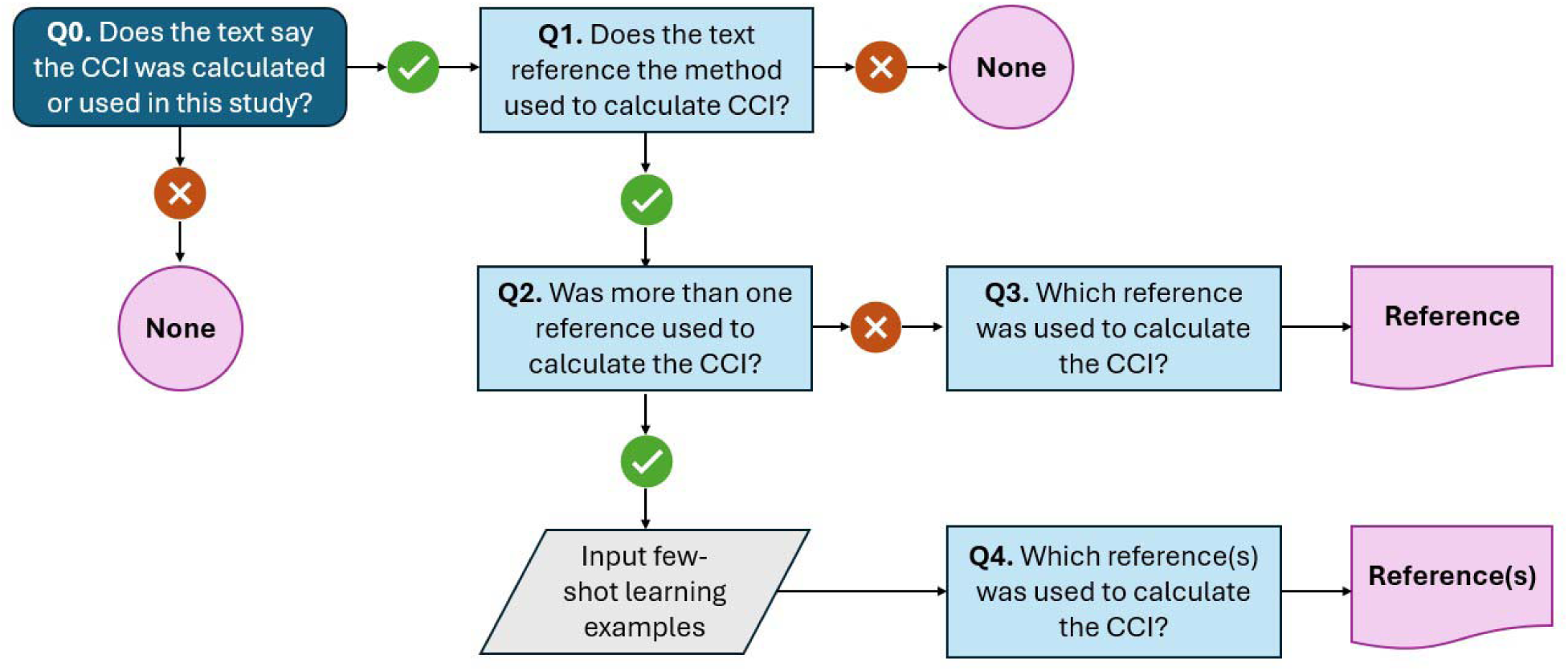
LLM prompt logic flowchart. Selected relevant paragraphs of text from the articles were given to the LLM along with the questions displayed. Positive (yes) or negative (no) responses from the LLM determine the flow through the flowchart. The full prompts for each question, as well as the few-shot learning examples, are given in Appendix A.

The next step in our analysis identified whether there is one or more than one reference that might indicate how the CCI was calculated. The identification and extraction of a single reference is more straightforward than identifying which of a set of references was used. This leads to the bifurcation shown after Q2 in Figure 2.

For paragraphs that were identified as having a single reference (Q2 = yes), we directly asked the LLM for the reference, while also providing an option to respond ‘None’ if the reference was ambiguous.

For paragraphs that had multiple possible references, we provided few-shot learning examples. Few-shot learning has been shown to increase the accuracy of LLMs^40–43^. We provided four examples with varying numbers of included and extracted references. We again provided the option to respond None if it was ambiguous which reference was used.

This data extraction workflow was implemented in Python using the Biopython^34^, pubmed_parser^36^, lxml^44^, numpy^45^, pandas^46,47^, and Beautiful Soup^48^ packages. We used the Llama 3.3 70B Instruct model^37^, though we expect other open-source models of similar development to perform similarly. To support more deterministic responses, we set the *temperature* parameter to 0 and the *top_p* parameter to 0.1. The code is publicly available in this GitHub Repo: https://github.com/NCTraCSIDSci/literature_analysis_with_llm^49^.

### Gold Standard Validation

To assess the performance of the model, we created a gold standard validation set by manually classifying a subset of articles across three outcomes: 1) articles with any number of extracted references, 2) articles where the LLM extracted fewer references as being used to calculate the CCI than were present in the article, and 3) articles where the LLM identified that the CCI was not calculated for use in that article. The first outcome provides validation on how well the LLM extracted references. The second and third outcomes provide validation on how well the LLM demonstrates improvement in large-scale literature review of previous methods such as string searching or reference matching.

Because model performance was expected to vary across the three outcomes, we calculated the necessary number of gold-standard classifications for each outcome individually. Using standard binomial precision methods with a 95% confidence level and 5% margin of error, we anticipated 90% accuracy for the first outcome requiring 139 classifications. We anticipated accuracies of 95% for the second and third outcomes, requiring 73 classifications each. The expected accuracies were informed by preliminary evaluations. Determining sample sizes individually in this way ensured that the validation was both feasible and appropriately rigorous across all outcomes^50,51^.

Two authors independently extracted results for each article; results that did not agree were settled by the review of a third author. These gold-standard results were then compared to the results produced by the LLM.

## Results

### Model Validation

We compared the results from the LLM-based method to our gold standard classifications, focusing on three categories of model performance.

First, we compared the results for 139 randomly selected articles. The extracted references agreed for 94.9% of the articles, indicating that extraction of references with the LLM produces results that are consistent with manual classification.

Second, we compared the results for 73 articles where the LLM extracted fewer references than were present in the article. The extracted references agreed for 94% of the articles.

Finally, to evaluate the rejection of articles that cited a CCI version but did not calculate it for use in that article, we compared the results for 73 articles. The gold standard classifications agreed that the articles did not calculate the CCI for 93% of the articles.

The second and third results demonstrate how an LLM-extraction routine can achieve results that are better than simple string searching or bibliographic matching. Both of these previous methods would have included references that are not actually used, biasing the results. The LLM routine can recognize nuances in articles that previous automated literature review methods struggle to achieve.

### LLM Data Extraction

Table 2 shows the overall results of the LLM data extraction process, displayed by year and CCI version.

**Table 2:**
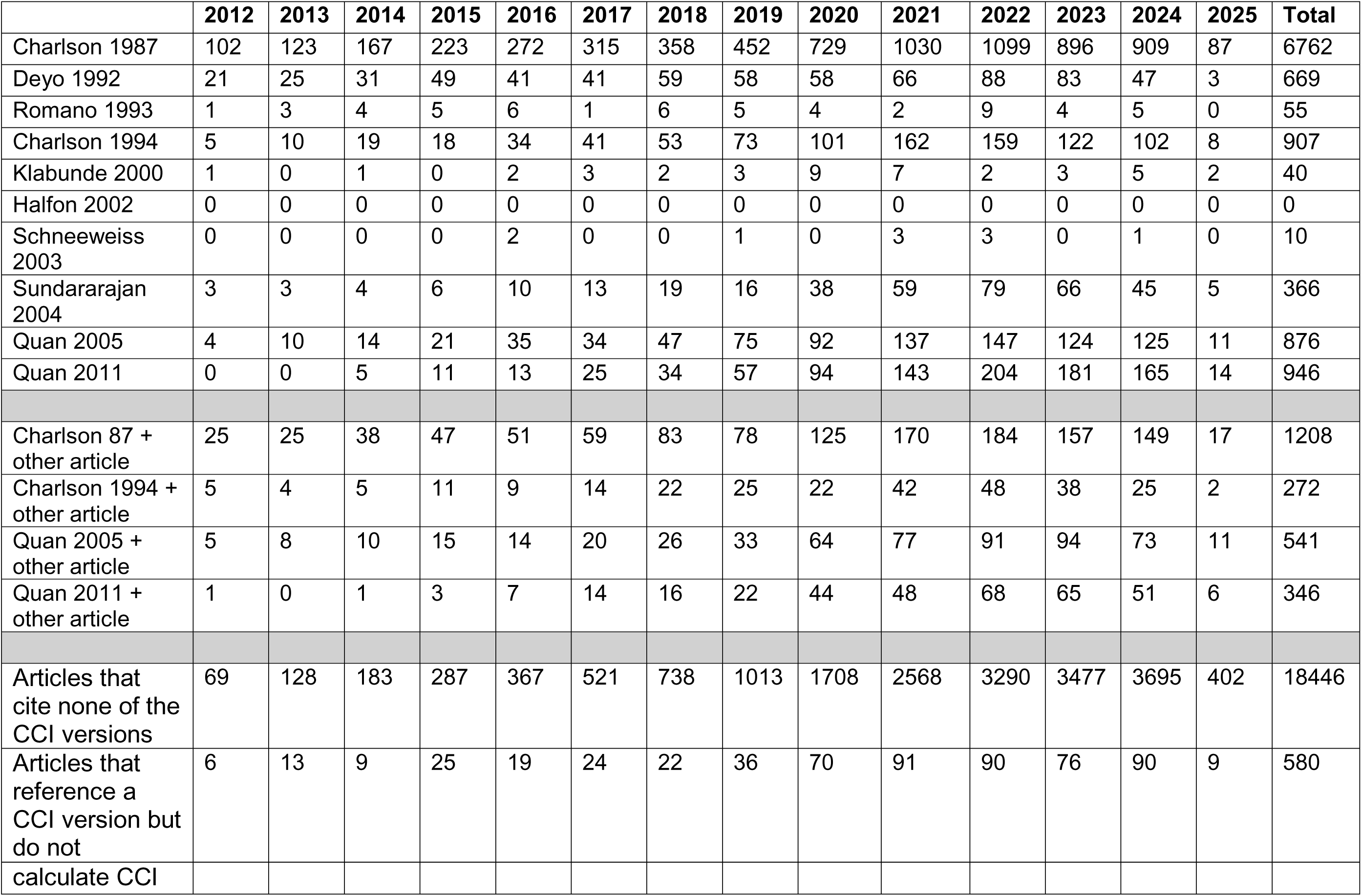
Number of extracted references that use that CCI version as a methodologic implementation per year. The top section of the table shows the number of times each CCI version was used as an individual implementation. The middle section of the table shows how often each of the top four papers from the first section was used in conjunction with another CCI version. The bottom section shows how often articles cited none of the CCI versions or cited a CCI version but did not calculate the CCI for use in the article.

### Articles Without References or That Did Not Calculate the CCI

Of the 31,767 articles included in this study, 58.0% (18,446) did not cite any of the CCI versions listed in 2. These articles did not proceed with the LLM data extraction routine. As such, we are not able to say definitively whether these articles solely discussed the CCI, calculated an individualized version of the CCI, or calculated one of the CCI versions in Table 1, but did not reference it.

There were 580 articles that contained references to at least one of the CCI versions listed in Table 1, but did not actually calculate the CCI for use in that article, as classified by our LLM data extraction routine. We emphasize this result, which highlights the ability of the LLM to discriminate between actual calculations of the CCI and, for example, general discussions about the CCI or explicit statements that the CCI was not calculated. String searches for references would not detect these articles, which account for nearly 4.5% of the articles analyzed.

### Articles with references

For the 13,030 articles that were determined to have calculated the CCI for implementation in that article, we divided the analysis into two portions. Figure 3 shows how often CCI versions were cited singularly. These are articles that cite only a single reference as used to calculate the CCI. We see that Charlson 1987 is overwhelmingly cited the most, with annual citations per year comprising ∼65% to ∼75% of the total.

**Figure 3:**
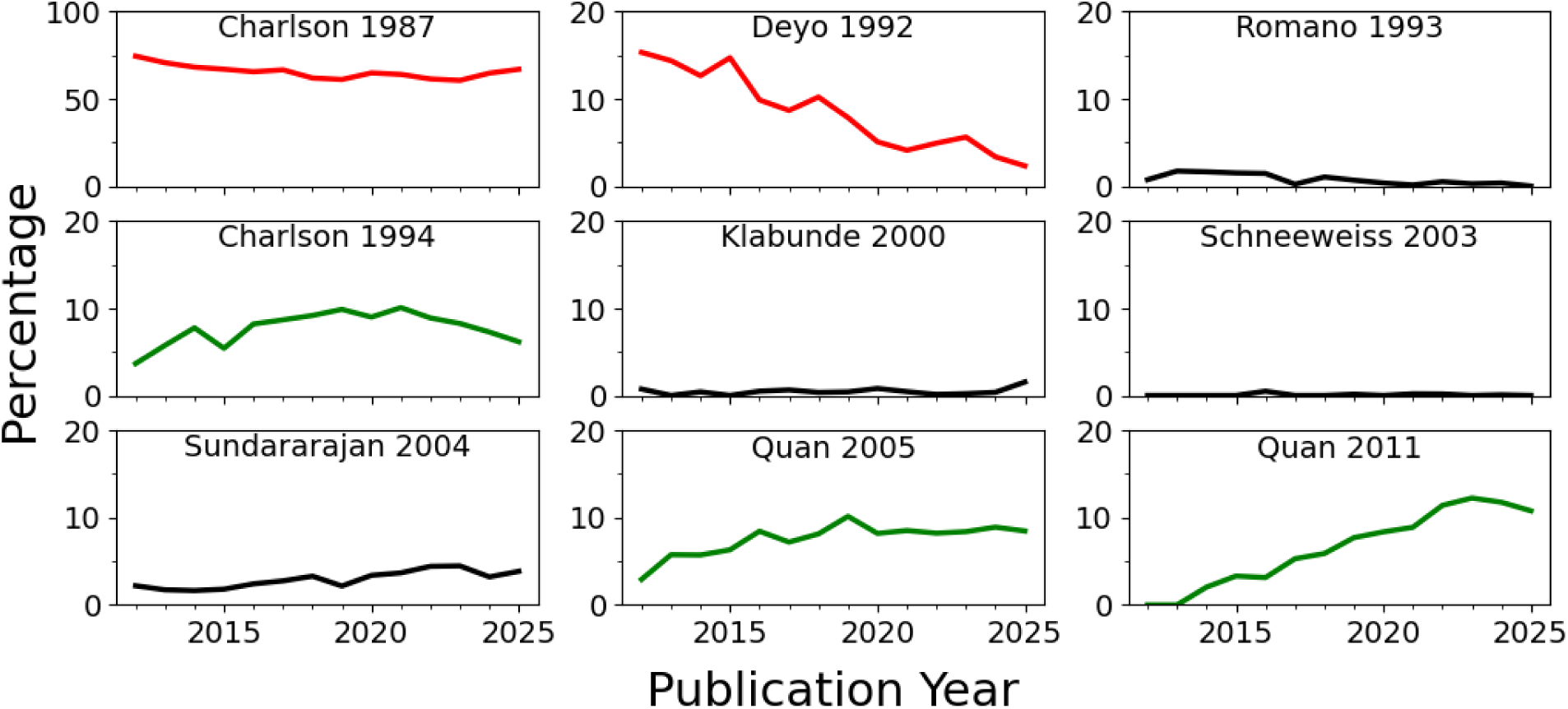
Single citations to individual CCI versions as a percentage of overall citations per year. Halfon, et al. (2002) is not shown because it was never cited by itself. Percentages are normalized by year to visualize relative changes of each. Charlson 1987 is on a different scale than all other versions. Lines in red indicate an overall decrease in percentage of citations per year, black indicates no significant change (+/-3%) in citations per year, and green indicates an increase in citations per year.

Charlson 1987 and Deyo 1992 both show overall decreases in the percentage of citations per year, with -8% and -13% changes, respectively. While the overall decrease in single citations to Charlson 1987 we consider a positive, it is still cited at a significantly higher rate than any other of the CCI we analyzed. Romano 1993, Klabunde 2000, Schneeweiss 2003, and Sundararajan 2004 show no significant change over time. Quan 2011, Charlson 1994, and Quan 2005 are cited and used with comparable frequency. All show moderate increase in the percentage of citations per year.

We then analyzed articles that used one or more than one version to calculate the CCI, as shown in the middle section of Table 2. The results are similar to those for single references, with Charlson 1987 being used most frequently. A total of 1,597 articles were determined to use more than one reference to calculate the CCI.

## Discussion

This study utilized a novel approach to characterize the evolution of CCI methods used in research over the last 10+ years of published literature. This type of study crucially allows assessment of CCI method utilization and evolution, as well as laying groundwork for future methods developments. Additionally, we demonstrated the feasibility and value of incorporating LLMs into literature review, which can facilitate accurate literature assessment at a scale not possible with human review.

There are several major lessons to learn from this study. Overall, Charlson 1987 has been and continues to be the most cited CCI version by a large margin, despite not being a version with an implementable method in the modern ICD-10 world. This finding would be reasonable if Charlson 1987 was cited along with another, modern CCI method, such as Quan 2005 but this dual citation occurs at only 15-20% of the rate of single citations annually (see Table 2).

Amongst subsequent, published method re-evaluations, a small number of papers represent the major evolution of CCI based on citations: Deyo 1992, Charlson 1994, Sundararajan 2004, Quan 2005, and Quan 2011. Moreso, despite the initial flurry of activity in considering the CCI from a methods standpoint, there does not appear to be a subsequent major methodologic evolution in CCI since Hude Quan’s papers 15+ years ago.

We have concerns about the prevalence of citations to Charlson 1987 in the literature without additional method citations. This suggests that the CCI has been somewhat ‘commoditized,’ representing a loose concept as opposed to a strict, validated method. One of the valuable aspects of the CCI is the accessibility of the method; however, this has potentially led to a lack of methodologic rigor over time. When the only citation in a modern paper for the CCI implementation is Charlson 1987, the research ultimately is opaque and non-reproducible. While the definition of methodological rigor may be subjective, transparency and reproducibility are not and only citing Charlson 1987 without additional clarification does not meet this standard. This concept of a ‘commoditized’ CCI extends beyond academic research, additionally, as private industry such as software companies and electronic health record (EHR) vendors also calculate the CCI without transparently sharing methods details. For instance, EPIC software calculates the LACE index^27^, which includes the CCI, but does not disclose in any known documentation the details of these calculations. This all suggests that the CCI has become a general concept, and researchers consider it sufficient to implement a method that ‘captures the spirit’ of the CCI without strictly using a validated method. The concern is that this lack of rigor degrades the value and interpretability of these scores particularly when applied over very large, broad patient populations, such as the entire patient population in a healthcare system EHR.

Ultimately, we advocate for two major focus areas within the research community employing the CCI. First, we advocate for a renewed methodologic focus with an acknowledgement that citing only Charlson 1987 is not sufficient when applying the CCI. Instead, the specific paper that describes the method employed should be cited in the research with any additional modifications to these methods described. The reader should be able to determine if a specific, published CCI method was employed, and either from that citation or from the methods text how comorbidities were identified, whether any additional comorbidities were added, the sources of comorbidities (particularly in EHR data which have multiple diagnosis code sources), and whether any statistical aspects were modified, such as category weights. Second, we advocate for a new methodologic assessment of the CCI in the real-world data era. In the modern era, there is a significant need to re-develop and re-validate the CCI using non-claims data sources commonly employed, particularly EHR data. Specifically, in standard EHR data, there is a need to investigate the utility and accuracy of the variety of discrete diagnosis inputs available (problem list, encounter, final billed, etc.) which do not exist in claims data^28^. Moreso, as common data models, particularly OMOP, become the primary form of EHR data researchers encounter there is a significant need for validating the CCI for optimal deployment in OMOP. The limited work on this so far has revealed issues and obstacles, and even the package provided by the Observational Health Data Sciences and Informatics (OHDSI) program for calculating CCI in OMOP data historically contains, likely unintended, errors of extra concept inclusion through broad inheritance that biases CCI results^29–31^. Finally, it would be valuable to evaluate what data standards are used to calculate the CCI (i.e. ICD-10 vs ICD-10-CM vs SNOMED) and compare the relative frequency of implementation of the Elixhauser Index, which has an annually updated list of comorbidity codes from AHRQ. The ongoing methods support and transparency of the Elixhauser Index with development specifically using the American ICD-10-CM terminology suggests Elixhauser may be a superior alternative to CCI in the modern era.

We demonstrated that using a LLM to extract information from research texts is an accurate, time-, and cost-efficient method for literature analysis. This method can not only be replicated with similar use-cases and LLM software but will also improve with the advancement of LLM engineering. Entire articles could be used, at increased cost, for more complicated data extraction.

This work demonstrates improvements over previous literature review methods, such as string searching or bibliometric reference extraction, with two main results:

1. The rejection of 580 articles that cited but did not actually calculate the CCI in the paper.
2. The 1,565 articles where the LLM extracted fewer references than the number included in the bibliography.

Previous methods of literature analysis would have missed both of these cases, which account for 16.4% of the total analyzed dataset. This demonstrates the ability of the LLM, with appropriately designed prompts, to detect and understand some of the nuance in this type of data extraction.

There are a few limitations to the methodologies employed in this study. Incorrectly or inconsistently formatted XML in articles might have led our preprocessing routine to not extract relevant paragraphs, though from testing we believe this to be the case for a very small number of articles. Additionally, if an article does not reference one of the CCI versions, we cannot tell whether the paper used a different version, created a specific implementation for that paper, or is ambiguous about how the CCI was calculated. Finally, we assumed that a description of if and how the CCI was calculated in a particular article would be present in the same paragraph as a reference to one of the major versions. We consider this a safe assumption, though we might have missed a small number of articles where this is not the case.

We introduced a novel method for conducting a characterizing review of a vast amount of published literature. We hope this enables future researchers to find trends and gaps in published research for similar use-cases. We have also identified a need for researchers to cite the methods used in the calculation of indices as well as for scientific publications and peer reviewers to ensure proper citation. Finally, a data standard is needed for managers of clinical data to readily provide documentation to researchers for all calculations performed in the dataset with appropriate citations.

## Data Availability

All data is publicly available as part of the PubMed Central Open Access Subset. The specific query is described in the Methods section.

## Code Availability

The code is publicly available in this GitHub Repo: https://github.com/NCTraCSIDSci/literature_analysis_with_llm

## Data Availability

All data produced in the present study are available upon reasonable request to the authors.

## Acknowledgements

We appreciate conversations with Jamie Conklin and Adam Dodd of UNC Libraries that assisted with the identification of the dataset.

The project described was supported by the National Center for Advancing Translational Sciences, National Institutes of Health, through Grant Award Number UM1TR004406. The content is solely the responsibility of the authors and does not necessarily represent the official views of the NIH. The funder played no role in study design, data collection, analysis and interpretation of data, or the writing of this manuscript.

## Author Contributions

JTF led the overall project, directing work, leading the programming and analysis, and writing the manuscript. CJ led the creation of the dataset and assisted with validation, figures, and writing. NF assisted with validation. PJL contributed to the conception and design of the work, feedback on methods, and writing. All authors reviewed and approved the final version of the manuscript.

## Competing Interests

The authors have no competing interests.

## Appendix A

Full text of the prompts used for the data extraction. See Figure 2.

### System Message 1

You are a literature review assistant. You will carefully review paragraphs and answer based on only what is present in the text. We are analyzing published literature to determine if it is clear which reference or references were used to calculate the Charlson Comorbidity Index (CCI) in this text. The text might include references that were not used to calculate the CCI, it might include just a single reference that was used, or it might contain multiple references that were used.

### Q0

You are an advanced language model tasked with analyzing a provided paragraph from a published research article. Your goal is to determine whether the text explicitly states that the Charlson Comorbidity Index (CCI) was calculated in the study. Please follow these guidelines: 1. Read the paragraph carefully and identify any mention of the Charlson Comorbidity Index. 2. Look for keywords and phrases that indicate calculation or use of the CCI, such as “calculated”, “assessed”, “used”, or similar terms. 3. If the paragraph suggests that the CCI was calculated or used in the analysis, respond with “Yes” 4. If the paragraph does not mention calculation or use of the CCI respond with “No” 5. Your response should be limited to “Yes” or “No” only, without any additional commentary or explanation. Please provide your response based on the analysis of the text provided. Don’t be strict.

### Q1

Answer Yes or No only. Does the following text reference which paper or weights was used to calculate the Charlson Comorbidity Index (CCI)?

### Q2

Answer Yes or No only. Does the following paragraph contain more than 1 reference to how the Charlson Comorbidity Index (CCI) was calculated?

### Q3

Which reference was used to calculate the CCI? Return only the reference as Last Name Year. If it is not clear which reference was used to calculate the CCI, return NONE.

### System Message 2 with few-shot learning examples

You are a literature review assistant. You will carefully review paragraphs and answer based on only what is present in the text. We are analyzing sentences from published literature to determine if it is clear which reference paper was used to calculate the Charlson Comorbidity Index (CCI) in this text. The following texts contain multiple references that might indicate how the Charlson Comorbidity Index (CCI) was calculated. We want to identify which reference or references were used to calculate the CCI in this paper. Sometimes these prompts will reference papers, but those were not used to calculate the CCI. Be strict.

Here is an example: Which reference or references were used to calculate the CCI in this paragraph? Return only the reference as Last Name and Year. If it is not clear what was implemented, return None. “In addition to DM, comorbidities defined by the Deyo’s Charlson Comorbidity Index [Deyo 1992] were examined using a revised mapping algorithm cited by Quan et al. [Quan 2005].” The correct response is Deyo 1992 and Quan 2005

Here is an example: Which reference or references were used to calculate the CCI in this paragraph? Return only the reference as Last Name and Year. If it is not clear what was implemented, return None. “The Charlson Comorbidity Index is sometimes used to measure somatic comorbidity in schizophrenic patients. The index includes 19 severe chronic disorders that are assigned a weighted score according to severity. The index was originally constructed to quantify the impact of comorbidity on mortality in a hospital setting among breast cancer patients (Charlson 1987) and later was adapted to ICD-10 diagnoses (Sundararajan 2004).” The correct response is Sundararajan 2004

Here is an example: Which reference or references were used to calculate the CCI in this paragraph? Return only the reference as Last Name and Year. If it is not clear what was implemented, return None. “We calculated the Charlson Comorbidity Index (Charlson 1987) using the (Quan 2005) implementation of ICD codes.” The correct response is Quan 2005

Here is an example: Which reference or references were used to calculate the CCI in this paragraph? Return only the reference as Last Name and Year. If it is not clear what was implemented, return None. “For most applications, the CCI score is calculated by manual record review or using claims data, typically coded using the International Classification of Diseases, 9th Version (ICD-9).(Deyo 1992) (Romano 1993) (Quan 2005) The former approach is costly and the latter introduces biases due to coding errors, heterogeneous coding conventions, and the granularity of the coding system.16 In addition, previous research has suggested that manually extracting comorbidity information from medical records is superior to the use of ICD-9 codes17 and claims data are not available until after discharge time.” The correct response is None

### Q4

Which reference or references were used to calculate or assess the CCI in this paragraph? Return only the reference as Last Name and Year. If it is not clear what was implemented, return None.

## Notes

### Competing Interest Statement

The authors have declared no competing interest.

### Funding Statement

The project described was supported by the (NCATS), National Institutes of Health, through Grant Award Number UM1TR004406. The content is solely the responsibility of the authors and does not necessarily represent the official views of the NIH.

### Summary of Updates

Gold standard validation expanded and clarified.

